# Abnormalities in migration of neural precursor cells in familial bipolar disorder

**DOI:** 10.1101/2021.04.22.21254208

**Authors:** Salil K. Sukumaran, Pradip Paul, Vishwesha Guttal, Bharath Holla, Alekhya Vemula, Harsimar Bhatt, Piyush Bisht, Kezia Mathew, Ravi K Nadella, Anu Mary Varghese, K Vijayalakshmi, Meera Purushottam, Sanjeev Jain, ADBS Consortium, Reeteka Sud, Biju Viswanath

## Abstract

Cellular migration is a ubiquitous feature that brings brain cells into appropriate spatial relationships over time; and it helps in the formation of a functional brain. We studied the migration patterns of induced pluripotent stem cell (IPSC)-derived neural precursor cells (NPCs) from individuals with familial bipolar disorder (BD), in comparison with healthy controls. The BD patients also had morphological brain abnormalities in magnetic resonance imaging. Time-lapse analysis of migrating cells was conducted, through which we were able to identify several parameters to be abnormal in cellular migration, including the speed and directionality of NPCs. We also performed transcriptomic analysis to probe the mechanisms behind aberrant cellular phenotype identified. Our analysis showed downregulation of a network of genes, centering on EGF/ERBB proteins. Present findings indicate that collective, systemic dysregulation may produce the aberrant cellular phenotype; which could contribute to the functional and structural changes in the brain, reported in bipolar disorder.

**SUMMARY STATEMENT:** We report abnormal cell migration patterns in neural precursors derived from bipolar disorder patients, which could contribute to already known structural changes in the brain.

## 1. Introduction

Bipolar disorder (BD) is a severe disabling psychiatric illness with a genetic basis, and neurodevelopmental origins [Gandal et al., 2018]. Many of the identified genes in BD risk are implicated in neurodevelopmental processes and variations in brain morphology [Mühleisen et al., 2018; Ganesh et al., 2019; Dai et al., 2020; Ithal et al., 2021]. Multiple studies have documented abnormalities in brain structure in BD [Magioncalda and Martino, 2021] including smaller brain size, reduced cortical gray and white matter [Ching et al., 2020], cortical thinning [Hibar et al., 2018], and decreased interneurons in the cerebral cortex and hippocampus [Harrison et al., 2020]. Cortical plasticity would also mediate the structural alterations and cognitive changes, seen over the life span in those with BD [Van Rheenen et al., 2020]. Such brain changes in BD have also been shown to be predicted by genetic risk [Abé et al., 2020].

One method to interrogate cellular alterations related to brain abnormalities is to study induced pluripotent stem cells (IPSCs) derived from patients, in whom changes have been detected on brain imaging. Although a direct link is difficult, this could help to identify potential contributory mechanisms to brain abnormalities. Previous IPSC-based studies have uncovered that BD pathogenesis is associated with differences in (a) expression of ion channels and membrane-bound receptors in neurons [Chen et al., 2014]; (b) neurogenesis and expression of genes of the WNT signaling [Madison et al., 2015]; and (c) mitochondrial abnormalities in patient-derived neurons [Mertens et al., 2015], as well as neural precursors [Paul et al., 2020; Osete et al., 2021].

Previous attempts to integrate human brain imaging with IPSC experiments in psychiatry [Johnstone et al., 2019; Vasistha et al., 2019] evaluated persons from multiple affected families with schizophrenia. Johnstone et al., (2019) showed reduction of cortical brain volumes with abnormal proliferation of neural precursor cells (NPCs), whereas Vasistha et al., (2019) showed oligodendrocyte proliferation and morphology deficits in individuals who had white matter alterations in the brain using diffusion tensor imaging. In addition, there is indirect evidence on the role of neuronal migration defects in BD [Uribe and Wix, 2012; Tabares-Seisdedos et al., 2006]. But there is no direct evidence on the migration defects that could contribute to bipolar disorder.

Here, we have performed a pilot study using IPSC-derived NPCs from a single family with two BD patients who had abnormal MRI scans. We have previously reported rare damaging variants related to cellular migration in these patients [Paul et al., 2020, supplementary table 1]. We hypothesized that there would be migration abnormalities in IPSC-derived NPCs of these patients. We find that both patient-derived NPCs displayed greater quasi-Brownian randomness in migration patterns, unlike the relatively directed movements in the control NPCs. Transcriptome analysis revealed expression changes in several genes known to regulate cellular migration, implicating the EGF/ERBB signaling pathway.

## 2. Results

### 2.1 Morphological changes observed on MRIs in brains of BD patients

In the scans of patient B1, we found that all tissue-specific brain volumes were below the 5th centile, demonstrating a clear deviation from age-related trends. In B2, lower gray matter (cortical at 10th centile and subcortical at ∼ 1/3rd centile) was detected, but the white matter volumes (∼50th centile) did not differ from age-related trends (Fig. 1A).

**Figure 1:**
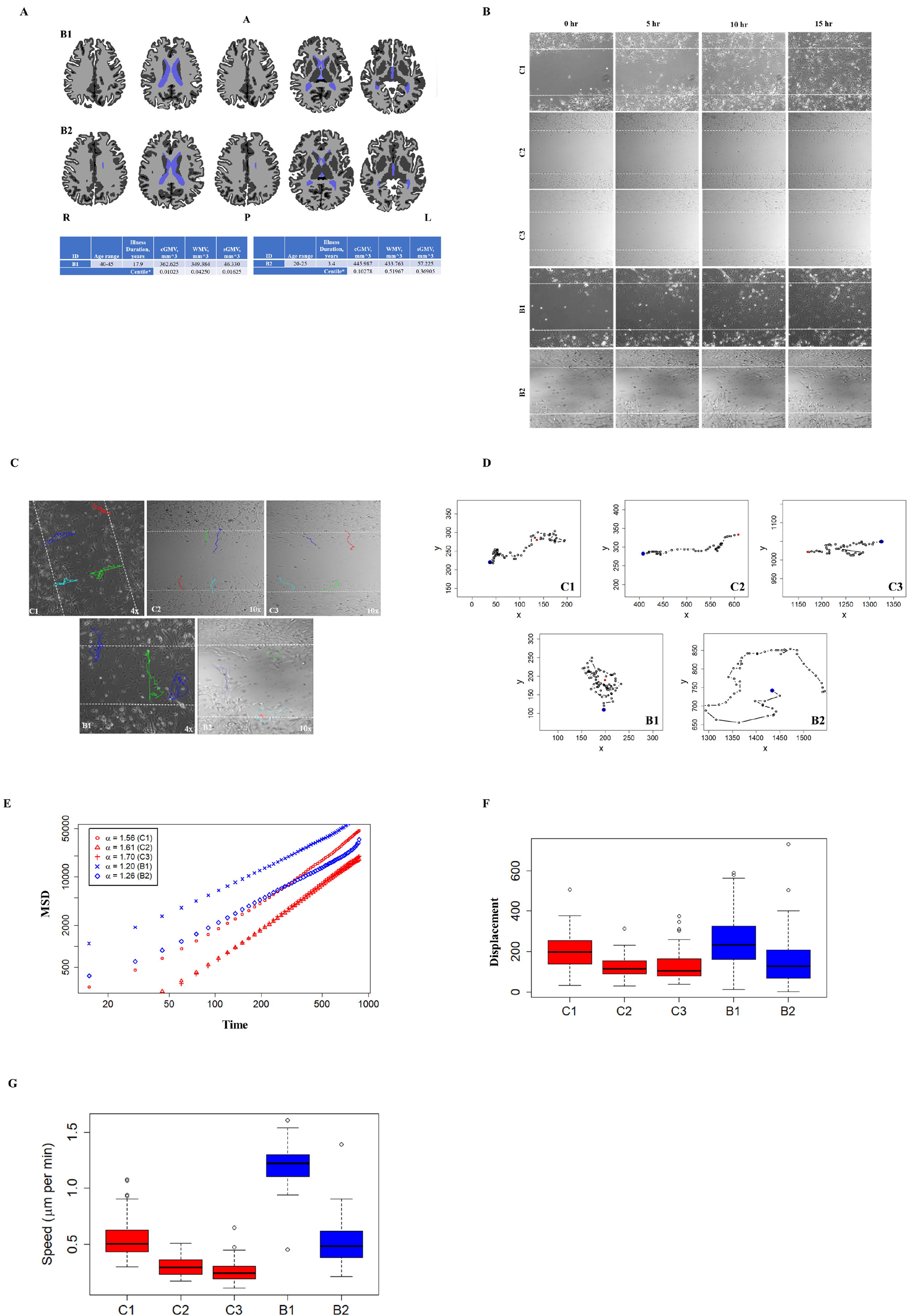
Abnormal migration phenotype in patient-derived neural precursors. A. Freesurfer based segmentation of T1w image into gray matter (dark gray), white matter (light gray) and ventricles (light blue). The top row shows axial slices for B1 and bottom row shows axial slices for B2. A, Anterior; P, Posterior; R, Right; and L, Left. cGMV, Total cortical gray matter volume; WMV, Total cerebral white matter volume; sGMV, Subcortical gray matter volume. B. Migration of neural precursor cells from C1-C3, B1, B2 at different time points. Only C1 closed the gap after 12 hours of migration. C. Direction of migration: C1-C3 migrated in a relatively directional pattern; B1 and B2 cells, on the other hand, migrated in a random, non-directed manner. Both BD lines showed back and forth movement, directional and circular patterns of migration. D. Sample migratory paths of control and patient derived lines over 15 hours. E. Mean squared displacement (MSD) as a function of time (in minutes), averaged over 100 cells per line. The value of MSD exponent a were 1.56 (with a 95% CI of 1.54 to 1.58), 1.61 (95% CI: 1.59 to 1.62) and 1.70 (95% CI: 1.69 to 1.71) for the three control lines, C1, C2, C3 respectively. In contrast, the MSD exponent a values were 1.20 (95% CI: 1.18 to 1.22) and 1.26 (95% CI: 1.24 to 1.28) for the B1 and B2 lines respectively. F. Box plots of displacement of the migrating cells after 15 hours (Mean ± SD; n = 100 cells per line). C1 moved a distance of 201.38 ± 81.78 µm, C2 a distance of 123.88± 50.34 µm, C3 a distance of 125.08 ± 67.39 µm, B1 a distance of 247.3±128.31 µm and B2 a distance of 150± 110.14 µm. G. Box plots of speed of migration for control and patient derived cells (Mean ± SD, over 15 hours; n = 100 cells per line): B1 cells migrated at a speed 1.21± 0.15 µm/min, B2 cells at 0.50 ± 0.17 µm/min; whereas C1 cells moved at 0.55± 0.17 µm/min, C2 cells at 0.3± 0.08 µm/min and C3 cells at 0.25± 0.08 µm/min. Mean speed of Control and B2 cell lines are statistically different from those of B1.

### 2.2 Aberrant cellular migration in patient-derived neural precursors

Cells from the control lines (C1, C2 and C3) migrated in a directed manner; while cells from the patient derived lines (B1 and B2) moved in a random (Brownian) manner throughout the duration of the experiment (Fig. 1B and 1C; Fig 1D for representative paths of cells). The patient-derived lines showed mixed patterns in the direction of migration: cells migrated in circular tracks as well as back-and-forth, in addition to some that migrated in a directed pattern (Fig. 1D; Supplementary Fig. S1). Quantitative analysis confirmed these visually observed differences between the migration trajectory of the controls and the patient-derived lines (Fig. 1E). The value of MSD exponent *α* were 1.56 (with a 95% confidence interval (CI) of 1.54 to 1.58), 1.61 (95% CI: 1.59 to 1.62) and 1.70 (95% CI: 1.69 to 1.71) for the three control lines, respectively. This indicates that control cells show a high degree of directionality with some amount of randomness. In contrast, the MSD exponent *α* values were 1.20 (95% CI: 1.18 to 1.22) and 1.26 (95% CI: 1.24 to 1.28) for the B1 and B2 lines respectively, suggesting a trajectory that was closer to a Brownian pattern of movement (Fig. 1E). Since the confidence intervals of the estimates of the MSD exponents of the control and patient-derived lines do not overlap, we conclude that the difference between the two groups is statistically significant. The overall displacement did not differ significantly (Fig. 1F). Additionally, though cells from B1 migrated at a faster speed, group-wise comparison indicated that migration speed did not differ significantly from cells derived from healthy controls (Fig. 1G).

### 2.3 Transcriptome analysis revealed differential expression of RNA transcripts related to cell migration

Our analysis for the migration-related genes (N=290) in the transcriptome data revealed that there were 61 transcripts in B1 and 58 in B2 that were differentially expressed, in comparison with C1. Of these, 29 transcripts were dysregulated across both patient-derived lines (Fig. 2B, C). The expression level of *LAMA1* and *NRG2* was confirmed using real-time q-PCR and was found to be downregulated (Supplementary. Fig. S3). Functional interaction of the above 29 using STRING highlighted a densely interconnected network of proteins, centered on ERBB proteins (Fig. 2D, Supplementary. Fig. S2).

**Figure 2:**
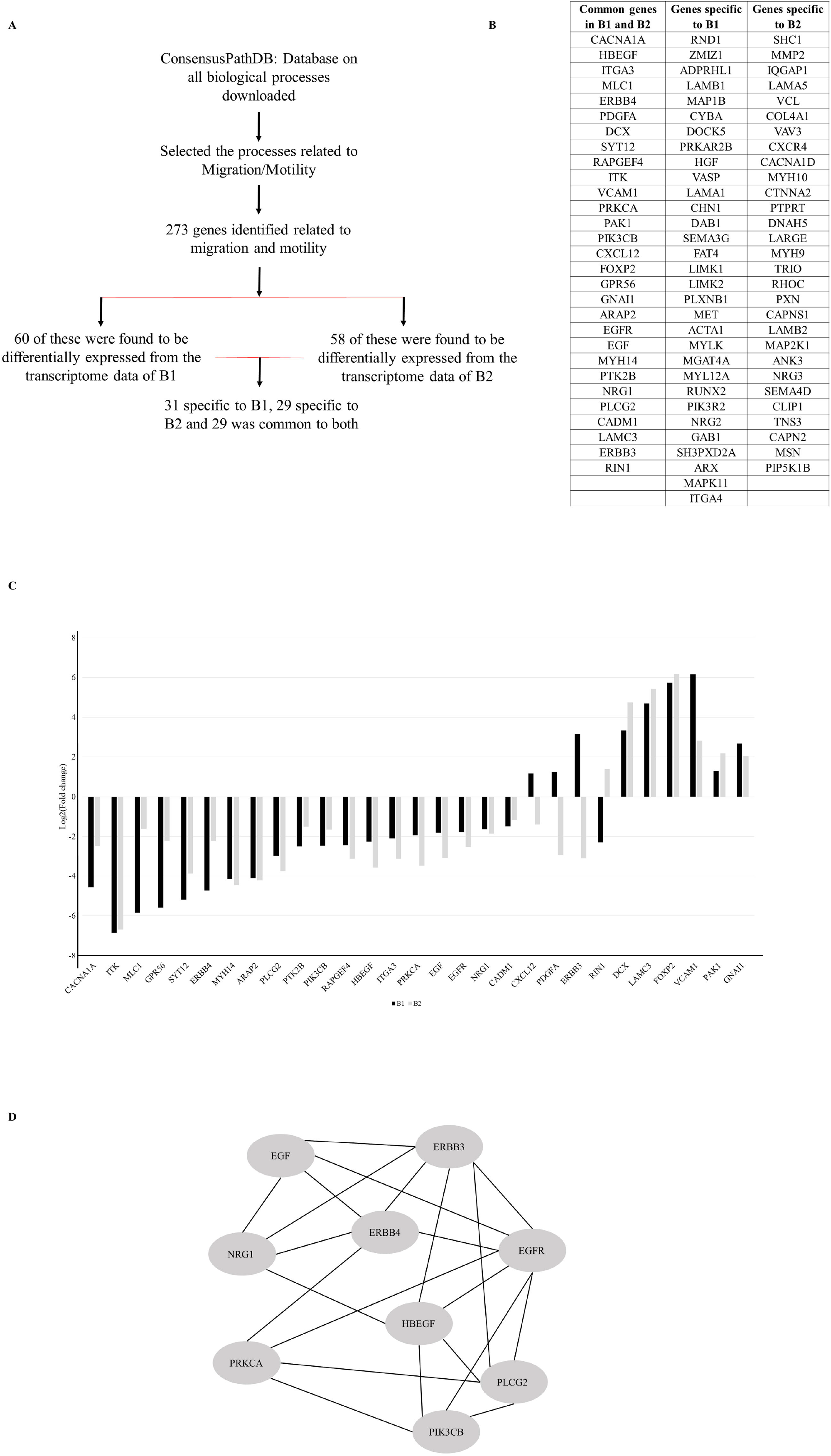
RNA-seq analysis shows dysregulated EGF/ERBB signaling pathway in patient-derived NPCs. A. Transcriptome analysis pipeline B. Transcripts common and unique to B1 and B2 patient-derived lines C. Comparison of fold change for common transcripts among the two patient-derived neural precursor lines. D. Protein interaction network, centered on ERBB4, that is downregulated in patient-derived NPCs. The interactions were plotted using STRING online database (https://string-db.org/). The entire interaction network is shown in Supplementary Figure S2.

## 3. Discussion

The BD patients in this study were chosen from a dense family, who had multiple rare damaging variants implicated in cellular migration, and had structural abnormalities noted on brain MRI (Fig. 1A). Cellular migration analysis showed that while patient-derived NPCs showed a random trajectory, the NPCs from healthy controls migrated towards other cells (directed/ ballistic movement), (Fig. 1C, D, E). Transcriptome analysis (Fig. 2A) showed that numerous migration-related genes were dysregulated in patient-derived NPCs (Fig. 2B, C).

The results of STRING analysis (Fig. 2D, Supplementary Fig. S2) revealed a network centered at ERBB proteins. This network includes several proteins including tyrosine protein kinases that function as cell surface receptors for neuregulins, EGF and other ligands. They are involved in organogenesis, including the brain, where they regulate many functions including cell proliferation, differentiation, migration and apoptosis. Proteins that are part of the EGF/ERBB network (Supplementary Fig. S2) fulfill different roles in migration of neural precursors. EGFR, PLCG2 and PRKCA, for instance, regulate migration via calcium signaling [Büttner et al., 2018]. Inhibition of ERBB receptors can lead to suppression of cancer cell migration [Momeny et al., 2017]; and inhibition of NRG1/ERBB2 signaling reduces migration of human glioma cells [Ritch et al., 2003].

The trajectory of migrating neural precursors lays down the foundations of developing central nervous system [Rahimi-Balaei et al., 2018; Barber et al., 2015]. The speed and direction of migrating cells can alter the regional cellular make-up, and therefore wiring of cortical areas. These observations at the level of individual cells may alter collective behavior (tissue organization); and is thus a question of fundamental importance in the context of brain disease, and behavior. Stochasticity is inherent in all biological systems, and its characterization may help us decipher local scale interactions among individuals, be it organisms or cells [Jhawar et al., 2020; Jhawar and Guttal., 2020; Bruckner et al., 2019]. An understanding of how single cells move, their inherent stochasticity and interaction can be crucial for understanding phenomena across scales, from tissue organization to wound healing and repair, and organization within the brain [Davidson et al., 2021; Otsuki and Brand, 2020; Zinner et al., 2020; Silva et al., 2019].

Overall, our previous work [Paul et al., 2020] and current analysis indicate that there are identifiable cellular abnormalities in NPCs derived from BD patients. These cells proliferate faster, and exhibit aberrant migration patterns. Whether these could, in part, be responsible for the deviations in gray and white matter noted on MRI imaging is a matter of conjecture at this point. It is also important to note that these findings are based on a small set of patients from one family, with absence of a family control. We further plan to conduct similar analysis in NPCs generated from a larger number of BD patients, and compare with not just population healthy controls, but also familial controls. Such experiments would be important to establish generalizability of present findings. The present work could also extend to an investigation in 3D organoids (mini-brains), to examine organizational aspects and functionality in more detail, currently in progress in our lab. The integration of deep clinical phenotyping with IPSC models in such sample sets will be crucial to understand the pathobiology of BD, and the mechanisms of recovery and resilience.

## 4. Materials and Methods

The NPCs from two BD-patients from a multiplex family, and three healthy control lines were used for experiments (pedigree and clinical details in Paul et al., 2020). Details of clinical assessments [Viswanath et al., 2018], magnetic resonance imaging (MRI) [Bhalerao et al., 2021; Holla et al., 2018; Parekh and Naik, 2021], generation of NPCs and their cellular characterization have been described earlier [Mukherjee et al., 2019; Paul et al., 2020]. The study was approved by the ethics committee of the National Institute of Mental Health and Neurosciences, Bengaluru, India, and conforms to the ethical norms and standards in the Declaration of Helsinki.

### 4.1 MRI analysis

Global brain volumes (gray and white matter) were calculated using the FreeSurfer software suite (v6.0). Individualized centiles for each tissue class were calculated using out of sample log likelihood estimation against the bootstrapped model parameters obtained from the expected age-related trends from a large aggregated database of reference brain volumes [Bethlehem R.A.I, 2021]. This database included normative brain volumes of Indian subjects for ages 6-60 years [Bethlehem R.A.I, 2021; Holla et al., 2020].

### 4.2 Assessment of migratory capacity of neural precursor cells

Cells were seeded in ibidi Culture-Insert 2-well in μ-dish (Cat. no. 81176, ibidi GmbH, Germany), at 85 - 90% confluency. Passage numbers for NPCs ranged between P10-P19. Movement of cells across the 500-μm gap in the ibidi dish was recorded in time-lapse images — every 15 min, for 15 hours with an Olympus microscope equipped with a camera (Hamamatsu) using a 10X dry objective. The experiments for each cell line were done in three biological replicates. Migration of NPCs were tracked for 12 hours, in a 37°C humidified chamber with 5% CO2, and quantified using ImageJ (Version IJ1.46r) with the plugin ‘Manual Tracking’.

### 4.3 Quantitative movement analysis of cellular migration

The paths of migrating NPCs were tracked with the help of X and Y coordinates from the images taken. From the trajectories of each cell, we estimated the (i) average speed (over 15 minutes intervals) and (ii) displacement between the initial and final locations (after 15 hours). The box-plot (Fig. 1F) shows displacement of 100 cells (cumulative from three experiments) for each cell line.

From (ii), we also computed mean-squared displacement (MSD) as a function of time separation between any two points along a cell’s trajectory. The MSD exponent α, which determines the functional relationship between MSD and the time lag (tau), given by *MSD*(*τ*) ∼ *τ*^*α*^, was also calculated. When *α* = 1, the cells are said to exhibit Brownian motion (or diffusive motion), whereas deviation from this value — termed anomalous diffusion values of *α* > 1 represent super-diffusive motion (or if *α* <1, sub-diffusive motion). A value of *α* = 2, implies that cells are migrating via a highly persistent or ballistic motion [Dieterich et al., 2008; Gal et al., 2013].

### 4.4 Transcriptome analysis

RNA-Seq of the NPCs was performed on the Illumina® Hi-Seq platform. Genes which showed >1-fold difference with FDR adjusted P-value < 0.05 were considered differentially expressed, as detailed elsewhere [Paul et al., 2020]. Since we observed migration deficits in patient-derived lines, we performed a targeted analysis focusing on genes already implicated in cellular migration. The gene list was extracted from the ConsensusPathDB analysis (Release 34) [Kamburov et al., 2011, 2009] (keywords “migration” and “motility”). In addition, the data from published references on genes related to migration were included [Snel et al., 2000; Suyama et al., 2003; Simpson et al., 2008; Wu et al., 2008; Bavamian et al., 2015; Madison et al., 2015; Buchsbaum and Cappello, 2019; Szklarczyk et al., 2019] to create a collated list of migration related genes (N=290).

### 4.5 Statistical tests

We wrote our own custom code in the statistical programming platform R to compute the speed, displacement, MSD and the MSD exponent. In addition, to compute the MSD exponent *α* and its confidence intervals, we performed a simple linear regression between log-transformed MSD and r, using the function *lm* in R. The slope of the resulting regression is an estimate of *α*. If the mean of the estimate of *α* of control lines do not overlap with the confidence intervals of *α* of the patient lines, we conclude that these two groups are statistically different with a p-value < 0.05.

We also performed two tailed t-test to compare migration speeds between control- and patient-derived NPC lines.

## Supporting information

C3 track path

B1 track path

B2 track path

C1 track path

C2 track path

## Data Availability

Available upon request

## Acknowledgements

The authors would like to thank Dr. Jitesh Jhawar, postdoctoral researcher, Department of Collective Behavior, Max Planck Institute of Animal Behavior, University of Konstanz, for his valuable suggestions on ImageJ analysis; Dr. Ravi Muddashetty and Dr. Dasaradhi Palakodeti for providing computational facilities for transcriptome analysis; Ms.Chitra B. and Mr. Mallappa M. for technical support. We are grateful to the participants and their families for their cooperation, as well as to clinicians and staff at NIMHANS for their assistance.

## Author Contributions

S.K.S - designed & performed NPC experiments, wrote the first draft, prepared figures, read and approved the final manuscript. S.K.S and P.P - Performed q-PCR. V.G. - Performed NPC quantitative movement data analysis. B.H. - MRI data analysis. A.V. and K.M. - Transcriptome data analysis. H.B. - Helped with the migration experiments. P.B. - Standardised the primers. R.K.N. - Performed clinical assessments. A.M.V and V.K. - Helped with the migration experiments. M.P. - Supervision of cellular experiments, read and approved the final manuscript. S.J. - Supervision of experiments, read and approved the final manuscript. R.S. - Supervision of cellular experiments, editing drafts, read and approved the final manuscript. B.V. - supervised clinical assessments and cellular experiments, read and approved the final manuscript. The overall study design was developed as part of the ADBS. All authors contributed to and have approved the final manuscript.

## Supplementary Methods

Total RNA was extracted using the Qiagen RNeasy Mini Kit (Cat. No: 74104). cDNA synthesis was performed with 1.5ug of RNA using High-Capacity cDNA Reverse Transcription Kit (Thermofisher Cat. No: 4368814). Sample cDNA and ‘no template control’ (NTC) were run in triplicates with the QuantStudio□6 Flex Real-time qPCR system (Thermofisher). The qPCR reaction was carried out with SYBR Green Master Mix (Takyon™ Low ROX) and 0.5 μM of each primer. Relative gene expression was estimated as previously described (Paul et al. 2020), and normalized to housekeeping gene *UBC*.

The primer sequences are:

*LAMA1* Forward : 5’ GAGCATGGAGAGATTCATACATC 3’

*LAMA1* Reverse : 5’ GGTCATGAGATCTGCATTGA 3’

*NRG2* Forward : 5’ GCAACGGCAGAAAGAACTCA 3’

*NRG2* Reverse : 5’ CTTCCCCAGGATGTTCTCGG 3’

*UBC* Forward : 5’ CTGGAAGATGGTCGTACCCTG 3’

*UBC* Reverse : 5’ GGTCTTGCCAGTGAGTGTCT 3’

## Supplementary Figures

**S1**. Representative recordings of cellular migration in C1-C3, B1, B2 lines. Migration tracks of individual cells were recorded by marking the position of the nucleus in individual frames. Colored lines indicate positions of individual cells during the duration of migration assay. While the migration pattern in C1-C3 is largely linear (S1-A, B, C), showing individual cells from either side of 500µm gap; in B1 and B2 lines (S1-D and S1-E), we encountered mixed patterns of cellular migration: apart from linear trajectory, these NPCs also demonstrated back-and-forth as well as circular motions. Images for migration of C1 and B1 cells were captured with 4x objective and C2, C3, B2 cell migration captured with 10x objective.

**S2**. Functional interactions among proteins shortlisted from Fig. 2C, explored using STRING online database. Only those proteins were included where direction of change of expression was similar in the two patient-derived lines.

**S3**. Real-time qPCR analysis of *LAMA1* and *NRG2* gene expression in B1 and B2 lines. Data are represented as Mean ± SD from three samples per individual group of two independent experiments.

**Supplementary Table 1:**

Rare damaging exome variants identified in patients.

