## Supplementary figures and images for "Abnormalities in migration of neural precursor cells in familial bipolar disorder"

### Supplimentary file S2

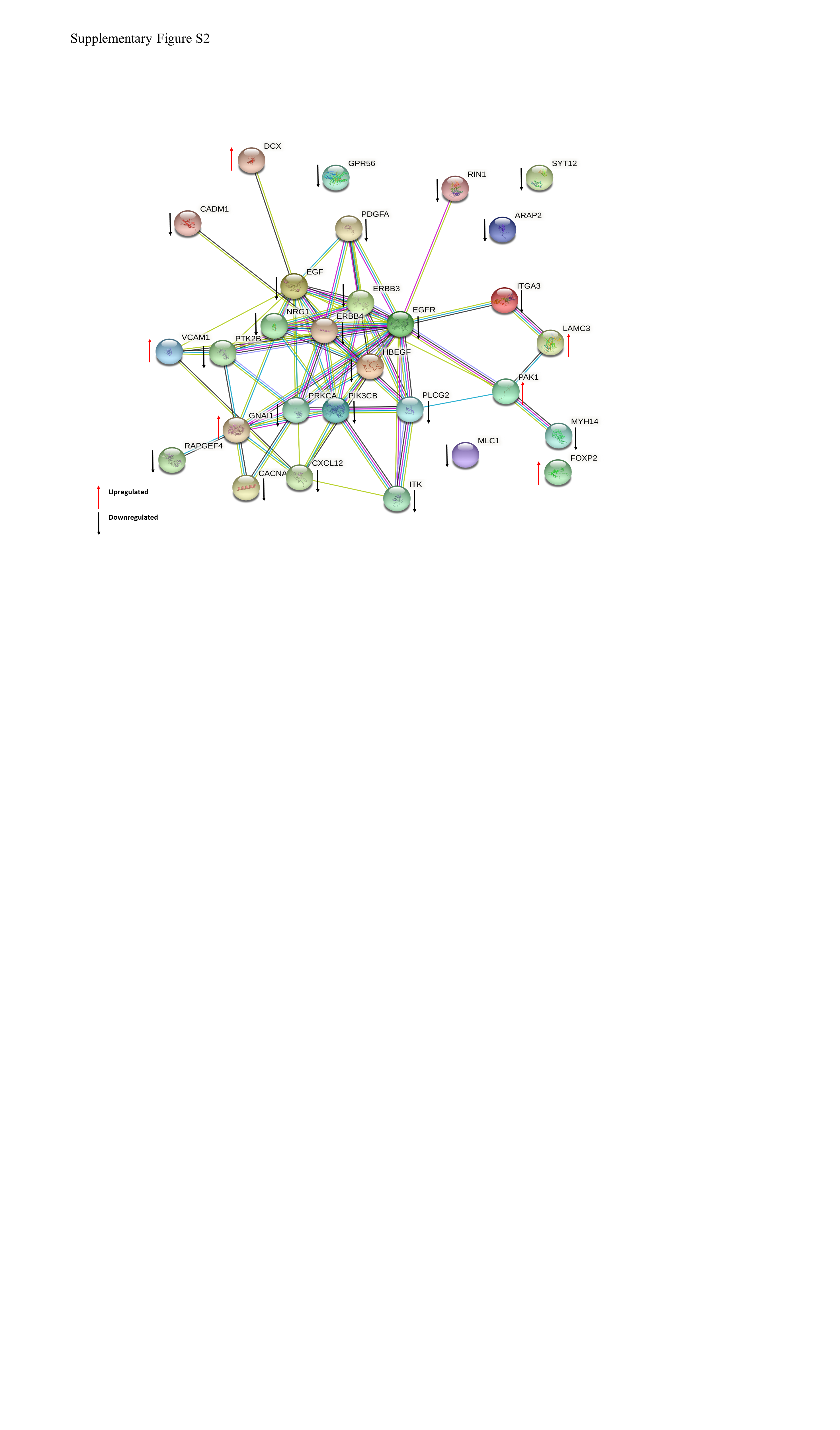

### Supplimentary file S3

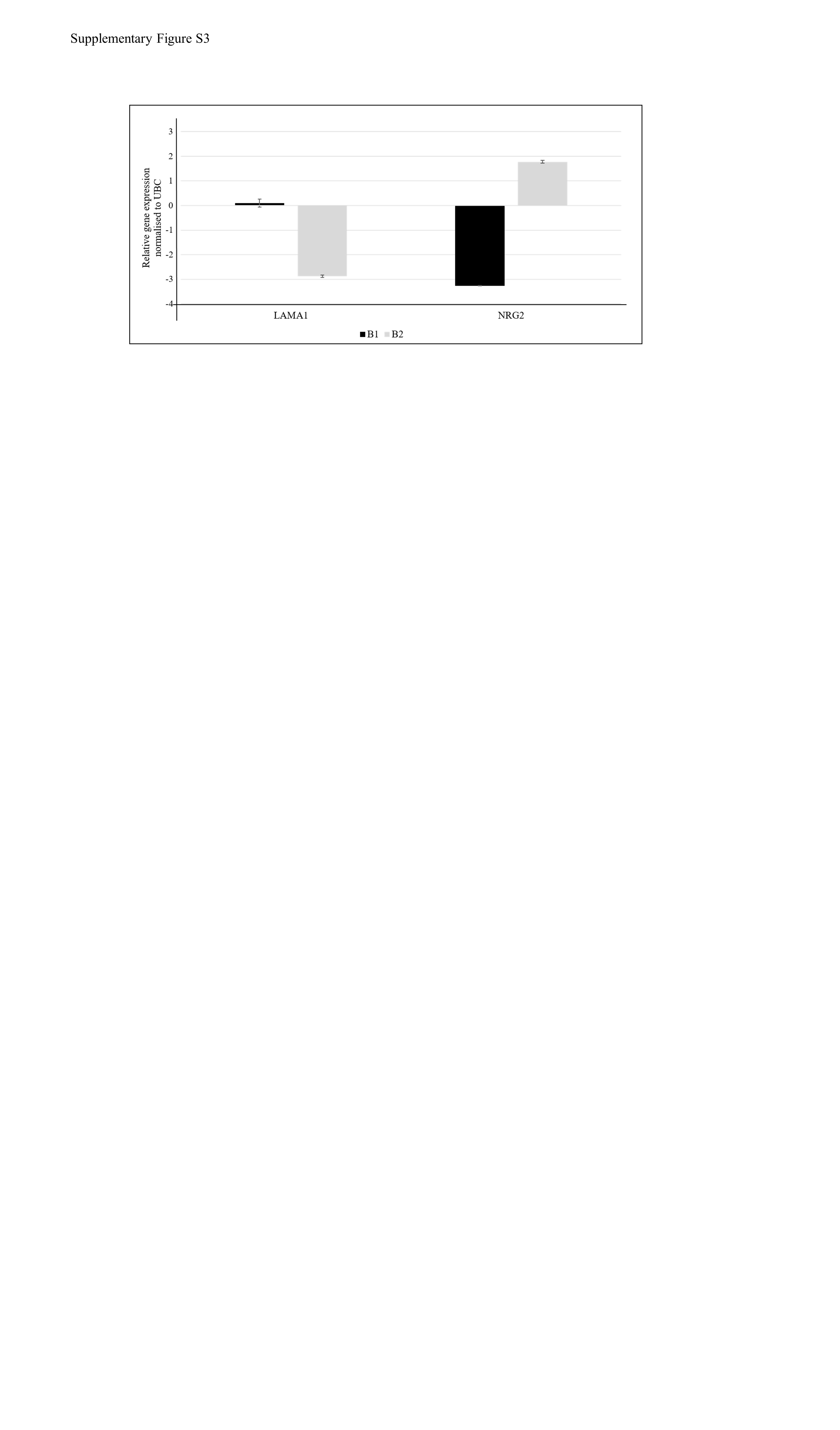
